# Exact Matching of Trabectome-Mediated Ab Interno Trabeculectomy to Conventional Trabeculectomy with Mitomycin C Followed for Two Years

**DOI:** 10.1101/2020.10.27.20165944

**Authors:** A. Strzalkowska, P. Strzalkowski, Y. Al Yousef, F. Grehn, J. Hillenkamp, N. A. Loewen

**Author notes:** **Correspondence to:** Nils Loewen, Department of Ophthalmology, University Hospital Würzburg, Josef-Schneider-Straße 11, 97080 Würzburg, Germany.

## Abstract

**Purpose:** We used exact matching for a highly balanced comparison of ab interno trabeculectomy (AIT) with the trabectome to trabeculectomy with mitomycin C (TRAB).

**Methods:** 5485 patients who underwent AIT were exact-matched to 196 TRAB patients by baseline intraocular pressure (IOP), number of glaucoma medications, and glaucoma type. Nearest-neighbor-matching was applied to age. Success was defined as a final IOP of less than 21 mmHg, IOP reduction of at least 20% reduction from baseline, and no secondary surgical interventions. Outcomes were measured at 1, 3, 6, 12, 18, and 24 months.

**Results:** 165 AIT could be matched to 165 TRAB. The mean baseline IOP was 22.3±5.6 mmHg, and the baseline number of glaucoma medications was 2.7±1.1 in both groups. At 24 months, IOP was reduced to 15.8±5.2 mmHg in AIT and 12.4±4.7 mmHg in TRAB. IOP was lower than baseline at all visits (p<0.01) and lower in TRAB than AIT (p<0.01). Glaucoma medications were reduced to 2.1 ± 1.3 in AIT and 0.2 ± 0.8 in TRAB. Compared to baseline, patients used fewer drops postoperatively (p<0.01) and more infrequently in TRAB than in AIT (p>0.01). Secondary surgical interventions had the highest impact on success and became necessary in 15 AIT and 59 TRAB patients. Thirty-two challenging events occurred in TRAB and none in AIT.

**Conclusion:** Both AIT and TRAB reduced IOP and medications. This reduction was more significant in TRAB but at the expense of four times as many secondary interventions.

**Key messages:** Despite vastly different IOP reduction and safety profile, ab interno trabeculectomy with the Trabectome and trabeculectomy with mitomycin C are both used as primary glaucoma surgeries. Exact matching allowed us to strictly focus on identical IOP and medications to create highly similar patient pairs for a balanced comparison that cannot be accomplished short of a randomized controlled trial. We found that trabeculectomy could achieve low IOPs and independence from drops, but trabeculectomies often required postoperative interventions. Trabectome patients had a lesser reduction of IOP and drops but needed far fewer interventions.

## Introduction

Trabeculectomy (TRAB), first performed in patients in 1961 by Sugar [1] and made more effective by the addition of mitomycin C in 1990 [2], continues to play a central role in surgical glaucoma treatment. However, postoperative challenges and complications can occur in up to 77-78% of patients [3–6]. This concern led to the development of non-penetrating and microincisional glaucoma surgeries (MIGS) [7]. One of the MIGS, ab interno trabeculectomy (AIT), is now often used both as a primary [8] and a secondary surgery [9] due to its proven, relative effectiveness in a range of glaucoma severity, including severe glaucoma [10–12]. AIT can also be performed after a failed trabeculectomy [13] or tube shunt [14]. This is surprising because TRAB allows aqueous humor to bypass the conventional outflow system, causing Schlemm’s canal and collector channels to atrophy [15, 16].

In theory, the IOP that AIT can achieve is limited by the episcleral venous pressure of 8 mmHg [17], yet most studies report IOPs around 15 mmHg. In contrast, TRAB can achieve very low IOPs, even below episcleral venous pressure, and eliminate drops [18, 19]. Hypotony [20, 21] is a feared consequence of too much pressure reduction. Phako-AIT has been compared to Phaco-TRAB in a randomized controlled trial but was stopped, with only 19 patients enrolled due to lack of clinical equipoise: TRAB was more complication prone than AIT, yet IOPs were similar [22]. There is scant data that directly compare TRAB to AIT. TRAB is a filtering surgery that drains aqueous humor into a subconjunctival reservoir, the bleb, while AIT enhances conventional outflow along its physiological route by removing outflow resistance at the level of the trabecular meshwork.

To address this unmet need, we applied an advanced method in statistics, *exact matching* [23–25], and created nearly identical pairs of AIT and TRAB based on IOP and number of topical glaucoma medications while using *nearest neighbor matching* for age. This design allowed us to compare the fate of highly similar eyes after being treated by these two distinctly different methods.

## Patients and Methods

### Study design

The study protocol was approved by the local ethics committee of the University of Würzburg, Germany, and performed in accordance with the ethical standards set forth in the 1964 Declaration of Helsinki and the Health Insurance Portability and Accountability Act. Because of its retrospective nature, informed consent was waived. All patients included in the study underwent either ab interno trabeculectomy using the Trabectome (AIT) or trabeculectomy with 0.02% mitomycin C (TRAB). An indication for surgery was an above-target IOP despite maximally tolerated topical treatment, as determined by a glaucoma specialist, or a need to reduce glaucoma medications despite stable IOP due to eye drop intolerance. We used data from the Trabectome Study Group database [13, 26] to increase the chances of an exact match to the TRAB group. The total number of cases available for an exact match was 5485. Patients younger than 20 years of age with neovascular or uveitic glaucoma were excluded. We recorded medical history, best-corrected visual acuity (BCVA [logMAR]), IOP (Goldmann tonometry [mmHg]), topical glaucoma medications, events leading to failure and serious, vision-threatening complications known from prior trabeculectomy and tube shunt studies, including aqueous misdirection, infection, wound leaks, hypotony maculopathy, choroidal effusions, choroidal hemorrhage, corneal decompensation. Success was defined as a final IOP ≤ 21 mm Hg, IOP reduction of at least 20% from baseline, and no secondary surgical interventions. Needling in the operating room in TRAB was counted as a surgical intervention. The decision to continue glaucoma drops was at the discretion of the treating specialist, as was the decision to advance to a different glaucoma surgery.

### Statistics

Data were described as frequency, percentage, mean±SD, median, and range. Continuous and categorical variables were compared with the Mann-Whitney U test and chi-squared test. Using *exact matching*, both groups were matched using preoperative IOP, glaucoma medications, type of glaucoma, and using *nearest neighbor matchin*g for age [27]. Each unit in AIT was matched using exact matching to all possible control units in TRAB. Whereas nearest neighbor matching selected the best match based on the distance to the value in AIT. P-values of less than 0.05 were considered statistically significant. Mean±SD was used to express continuous variables. Statistical analyses were performed using R [28]. A Kaplan-Meier plot was generated.

### Surgical technique

AIT was performed as described before [29]. Briefly, a 1.6 mm uniplanar, temporal clear corneal incision was created. Under direct gonioscopic visualization, the tip of the Trabectome handpiece (MST MicroSurgical Technology, Redmond, WA, USA) was inserted into Schlemm’s canal. The TM was ablated counterclockwise, followed by clockwise ablation with a total length of about 120 degrees [30–32]. Ablation was started with the power set to 0.8 mW and increased as necessary. The handpiece was withdrawn from the anterior chamber.

In TRAB, the eye was rotated downward with a traction suture, as described before [33]. A 5 mm limbus-based peritomy was created at the anatomic 12 o’clock position. A sub-tenon pocket was fashioned to accommodate a sponge soaked with mitomycin C (MMC) at a concentration of 0.2 mg/ml for 3 minutes. A 3.5 mm x 4 mm scleral flap was created. A 0.8 mm x 2 mm sclerotrabecular block was excised to enter the anterior chamber. A peripheral iridectomy was made. The scleral flap was secured with 10-0 nylon to allow visible percolation of aqueous, and the conjunctiva was closed with an interlocking running suture resulting in a diffusely forming bleb. All patients received daily subconjunctival injections of 2 mg of 5 fluorouracil (5-FU) for one week except when IOP was at or below 5 mmHg or the Seidel test was positive for a bleb leak.

In both AIT and TRAB, postoperative treatment consisted of a combination of a topical antibiotic for one week and a steroid tapered over four weeks. Glaucoma medications were stopped on the day of surgery and restarted as needed.

## Results

Using exact *matching*, 165 AIT were paired with 165 TRAB. The baseline IOP and glaucoma medications in AIT and TRAB were identical. IOP was 22.3±5.6 mmHg, and medications were 2.7±1.1. The demographic characteristics are presented in **Table 1**. AIT patients had an average age of 50±15 years, significantly younger than TRAB 67±11 (p<0.01). Despite their younger age, 60% of AIT patients had prior surgeries, many more than TRAB, with only one prior trabeculectomy (0.6%). Prior procedures in AIT included 13 (8%) ALT, 38 (23%) SLT, 20 (12%) cataract surgeries, 9 (5%) other surgeries, as well as major glaucoma surgeries consisting of 16 (10%) trabeculectomies and 4 (2%) tube shunts. Forty-six (28%) AIT operations were combined with phacoemulsification.

**Table 1.** Demographics (*=p<0.05).

|  | AIT<br>n = 165 | TRAB<br>n = 165 |
| --- | --- | --- |
| **Age\*** |  |  |
| Mean±SD | 50±15 | 67±11 |
| Range | (16, 87) | (25, 93) |
| **Gender** |  |  |
| Female | 72 (44%) | 86 (52%) |
| Male | 92 (56%) | 79 (48%) |
| **Diagnosis** |  |  |
| Primary open angle glaucoma | 130 (79%) | 130 (79%) |
| Pseudoexfoliation glaucoma | 32 (19%) | 32 (19%) |
| Pigment dispersion glaucoma | 3 (2%) | 3 (2%) |
| **Prior Procedures and Surgeries\*** |  |  |
| Argon laser trabeculoplasty (ALT) | 13 (8%) | ----- |
| Selective laser trabeculoplasty (SLT) | 38 (23%) | ----- |
| Cataract | 20 (12%) | ----- |
| Trabeculectomy | 16 (10%) | 1 (0.6%) |
| Shunt | 4 (2%) | ----- |
| Other | 9 (5%) | ----- |
| **Combined Surgeries\*** |  |  |
| Combined with phaco | 46 (28%) | ----- |
| Combined with other | 4 (2%) | ----- |

At one month, IOP decreased to 16.7±5.6 in AIT and 11.1±4.0 in TRAB (**Table 2**; **Figure 1a**, all-time points p<0.01). IOP stayed relatively similar throughout the study, trending towards 15.8±5.2 mmHg in AIT and 12.4±4.7 in TRAB 24 months. Medications had declined from 2.7±1.1 in each group to 2.4±1.5 in AIT and 0.0±0.2 in TRAB at one month. At 24 months, they were at 2.2±1.3 in AIT and 0.2±0.8 in TRAB (all time points p<0.01). AIT had a significantly higher IOP and number of medications throughout the study (p<0.01). In TRAB, the medication reduction was significant (p<0.01, **Table 2; Figure 1b**) at all time points of the study compared to baseline. In AIT, the medication reduction was small and only became statistically significant after three months (after three months p<0.05).

**Table 2.** Mean IOP and number of Medication for AIT and TRAB at each follow-up time point (Welch two-sample t-test, significance level set at ≤0.05)

| Time | IOP |  |  | Rx |  |  |
| --- | --- | --- | --- | --- | --- | --- |
|  | AIT | TRAB | P-value | AIT | TRAB | P-value |
| baseline | 22.3±5.6 | 22.3±5.6 |  | 2.7±1.1 | 2.7±1.1 |  |
| 1 month | 16.7±5.6 | 11.1±4.0 | <0.01* | 2.4±1.5 | 0.0±0.2 | <0.01* |
| 3 months | 16.0±3.9 | 11.4±3.9 | <0.01* | 2.3±1.5 | 0.0±0.3 | <0.01* |
| 6 months | 16.3±4.1 | 11.5±3.4 | <0.01* | 2.1±1.4 | 0.0±0.4 | <0.01* |
| 12 months | 16.0±3.9 | 11.9±3.3 | <0.01* | 2.1±1.3 | 0.1±0.4 | <0.01* |
| 18 months | 15.3±3.4 | 12.1±3.4 | <0.01* | 2.2±1.4 | 0.1±0.5 | <0.01* |
| 24 months | 15.8±5.2 | 12.4±4.7 | <0.01* | 2.2±1.3 | 0.2±0.8 | <0.01* |

**Figure 1.**
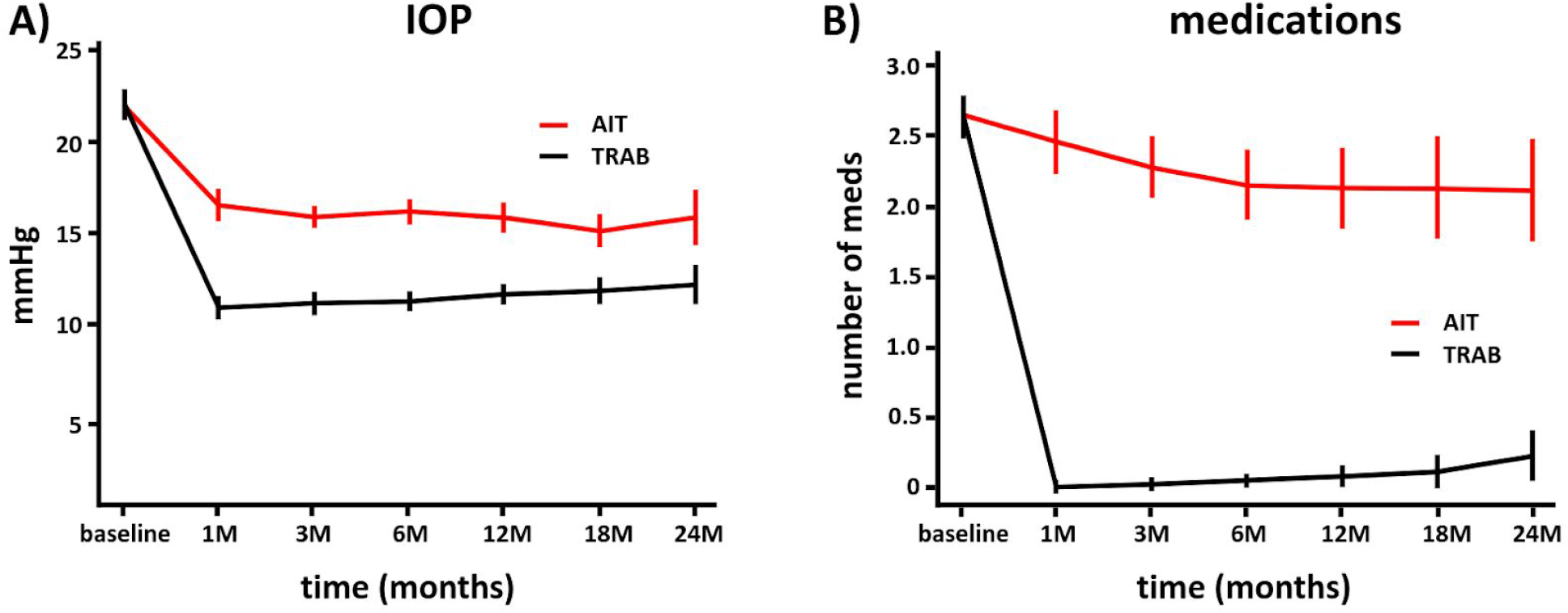
IOP and number of medications for AIT and TRAB. A) IOP decreased to a relatively stable level in AIT and slightly trended up in TRAB. B) Glaucoma medications continued to decrease for 6 months and remained at this level and slightly trended up in TRAB after a sharp drop at 1 month. (mean±SEM; subject count at each time point).

When all interventions were counted towards surgical failure, success was achieved in 93% of AIT and 58% of TRAB eyes (difference p<0.01, Supplementary fig. 1). Additional surgery was needed in 15 (9%) of AIT and 59 (36%) of TRAB. In TRAB, 24 (14.9%) patients needed bleb needling, often considered a minor secondary surgical intervention, 33 (20.0%) an open bleb revision, one iris repositioning (0.6%) and one cyclodestruction (0.6%). In AIT, 15 of 18 patients with high IOP received a tube shunt. Success criteria were not fulfilled in 18 AIT (11%) and one TRAB patient (0.6%, **Table 3)**. In TRAB, 17 (10%) eyes had a shallow anterior chamber, 16 (10%) developed a cataract, 7 (4%) choroidal effusion, 6 (4%) with bleb leaks, and 3 (2%) with iris adhesion.

**Table 3.** Number of intervention for AIT and TRAB within 24 months.

| Group | AIT | TRAB |
| --- | --- | --- |
| Patients with failure | 33 (20%) | 60 (36.6%) |
| due to intervention including bleb needling | 15 (9%) | 59 (36%) |
| due to intervention excluding bleb needling | 15 (9%) | 35 (14.9%) |
| due to high IOP | 18 (11%) | 1 (0.6%) |

An additional intervention was needed in 15 (9%; 45% of all failures) of AIT and 59 (36%; 98% of all interventions) of TRAB (Table 3). In AIT, 15 (9%) patients (45% of all interventions) received a tube shunt. In TRAB, one patient needed a secondary glaucoma surgery. However, 24 (14.9%) TRAB patients (41% of all interventions) needed bleb needling in the operating room, 33 eyes (20.0%, 56% all interventions) needed an open scleral flap revision most of which resulted in successful IOP control, one cyclodestruction (0.6%, 1,5% of all failures) and one iris repositioning (0.6%, 1,5% of all failures). 17 (10%) TRAB eyes had a shallow anterior chamber, 16 (10%) developed a cataract, 7 (4%) choroidal effusion, 6 (4%) bleb leaks, and 3 (2%) iris adhesion.

Failure to control IOP (final IOP ≤ 21 mm Hg, IOP reduction of at least 20% from baseline) was more common in AIT, where it occurred in 18 (10.9%) compared to only one (0.6%) in TRAB.

## Discussion

This study aimed to compare IOP and glaucoma medication reduction by two distinctly different glaucoma surgeries, AIT and TRAB, commonly used as primary surgical treatments. We wanted this comparison to be agnostic to disease severity and instead generate a highly balanced comparison, strictly focusing on IOP and medications in our exact matching algorithm. Matching is a nonparametric method designed to control confounding [34]. Propensity score matching would be well suited to compare such dissimilar datasets [35, 36], while exact matching is more commonly used to compare similar data [37, 38]. However, exact matching allowed us in this study to more carefully examine two of the most important variables in glaucoma care, IOP, and eye drops needed to achieve this IOP.

Techniques and mechanisms of IOP reduction of AIT and TRAB are vastly different. TRAB lowers IOP by allowing aqueous humor to bypass the conventional outflow system and drain into a bleb, a sub-tenon reservoir, from which aqueous is removed by lymphatics, absorption, and transconjunctival diffusion [39]. It is well established [40, 41] that trabeculectomies can lower IOP and reduce medications profoundly, but have a higher risk of complications and often need additional interventions. In contrast to that, AIT removes a primary substrate of outflow resistance, the trabecular meshwork [29]. Hypotony related complications do not normally occur in AIT because the IOP cannot be lower than the episcleral venous pressure. Recent studies have demonstrated that the average IOP after AIT is higher than one would expect based on the 8 mmHg present in the episcleral veins [42]. There is new evidence of a post-trabecular outflow resistance [43–45] that could cause AIT to fail but would not affect the results of TRAB.

Trabeculectomy is known to increase the cataract progression rate to up to twice that of controls [46, 47]. In our study, TRAB had a higher rate of cataract progression than AIT. AIT patients were younger, 20 had already had cataract surgery, and 46 had cataract removal combined with AIT. It is possible that trabeculectomy surgeons focused on IOP and avoided the additional challenges and risks of cataract surgery in the same session. Combining cataract surgery with AIT does not contribute to IOP reduction, however [48]. In contrast, cataract surgery can lower IOP by 1.5-3 mmHg when done on its own [49–51] or when combined with a trabecular bypass stent [52], due to a trabeculoplasty-like effect [37, 53]. However, too much trabecular meshwork is removed in AIT to be affected by this, and more circumferential flow is generated compared to trabecular bypass stents [54].

Prior laser procedures and incisional glaucoma surgeries can jeopardize the success of AIT. AIT patients had a considerably higher rate of prior surgeries, putting them at a disadvantage in our study. Argon laser trabeculoplasty (ALT) causes coagulative damage to the trabecular meshwork and Schlemm’s canal [55]. These create perceptible stops during AIT that likely limit circumferential flow. Selective laser trabeculoplasty (SLT) does not do that [56].

Consequently, when SLT fails to lower IOP, it could indicate that the distal outflow system is dysfunctional, reducing the chances that AIT will work. This theoretical consideration was not seen in Klamann et al.’s study, however [57]. A failed prior tube shunt or trabeculectomy may also reduce chances of a successful AIT as there is histological evidence that Schlemm’s canal and collector channels atrophy [15, 16]. This concern did equally not materialize in two studies that found that there was a reasonable IOP reduction by AIT after failed tubes [14] and trabeculectomies [13].

TRAB patients had a significantly better reduction of IOP and medications at two years in our study, whereas patients in AIT still required more than two glaucoma drops on average. While 15 (9%) of patients in AIT needed another major glaucoma operation (tube shunt), there was only one further glaucoma procedure (cyclophotocoagulation) in TRAB. This advantage of TRAB comes at the price of more intense postoperative care (i.e., postoperative 5-fluorouracil injections) and additional interventions such as bleb needling and sometimes surgical revisions. However, many of these interventions, albeit cumbersome for the patient and the surgeon, often have no negative impact on the final outcome. In addition, postoperative challenges such as choroidal effusion and a shallow anterior chamber often resolve on their own.

Although our AIT patients were younger, 28% had concurrent cataract surgery, and 12% were already pseudophakic. None of these conditions would generate a better IOP reduction, as we have shown before [42, 48]. The AIT data used in our study to create exact matches do not represent the experience of a single surgeon or single center but represents the average of multiple centers and surgeons. Surgeons had to submit their first cases as part of a post-market surveillance requirement by the US Food and Drug Administration (FDA) [13, 26]. Likely, most AIT surgeons were still on the learning curve, ranging from 5 to 30 eyes, as observed in an AIT training model [58].

Limitations of our study include its retrospective nature. This shortcoming is countered by *exact matching*, a robust automated statistical method that creates a highly balanced comparison and reduces confounding [59, 60]. Although the algorithm discarded AIT data, we were fortunate to be able to afford this and create identical IOP and glaucoma eye drop matches with the much more limited number of TRAB so that only 31 were lost. Had we increased the number of matching variables to improve the precision of matching, many more patients would have been excluded, reducing the study sample size and variability of the patient population [60, 61]. Consequently, we applied nearest neighbor matching to age, and AIT patients ended up with a younger average age of 50±15 years compared to TRAB with 67±11 years. Although this age difference is considerable, it would have put AIT only at a slight disadvantage of approximately 0.5 mmHg as predicted by our IOP calculator [42].

Another limitation is that we ignored the individual target IOP to focus on IOP and medications. Using a common definition of success, which considered any secondary surgical intervention [62, 63], AIT had a higher success rate than TRAB. However, many trabeculectomists would not consider needling or revisions a complication as long as the target pressure is achieved eventually. Our TRAB achieved an IOP reduction equal to that in the TVT study but with fewer medications, while our AIT appeared to be similar to the tubes in that study [64].

Our study confirmed that TRAB remains a good choice for patients with low IOP targets and need to reduce drops. This procedure requires patients to accept a high chance of postoperative challenges. Analysis of AIT patients matched to the relatively low preoperative IOP of TRAB patients confirm that AIT is a low-risk procedure that achieves physiological IOPs. Fortunately, TRAB and AIT do not exclude each other as AIT can be performed after TRAB has failed [13] and vice versa [65].

## Data Availability

All data is contained in the manuscript.

## Compliance with Ethical Standard

### Ethical approval

All procedures performed in studies involving human participants were in accordance with the ethical standards of the University of Würzburg and with the 1964 Helsinki declaration and its later amendments or comparable ethical standards.

### Funding

No funding was received for this research.

### Potential Conflict of Interest

The authors certify that they have no affiliations with or involvement in any organization or entity with any financial interest (such as honoraria; educational grants; participation in speakers’ bureaus; membership, employment, consultancies, stock ownership, or other equity interest; and expert testimony or patent-licensing arrangements), or non-financial interest (such as personal or professional relationships, affiliations, knowledge or beliefs) in the subject matter or materials discussed in this manuscript.

### Informed consent

This type of study does not require informed consent.

---

**Supplemental figure 1.**
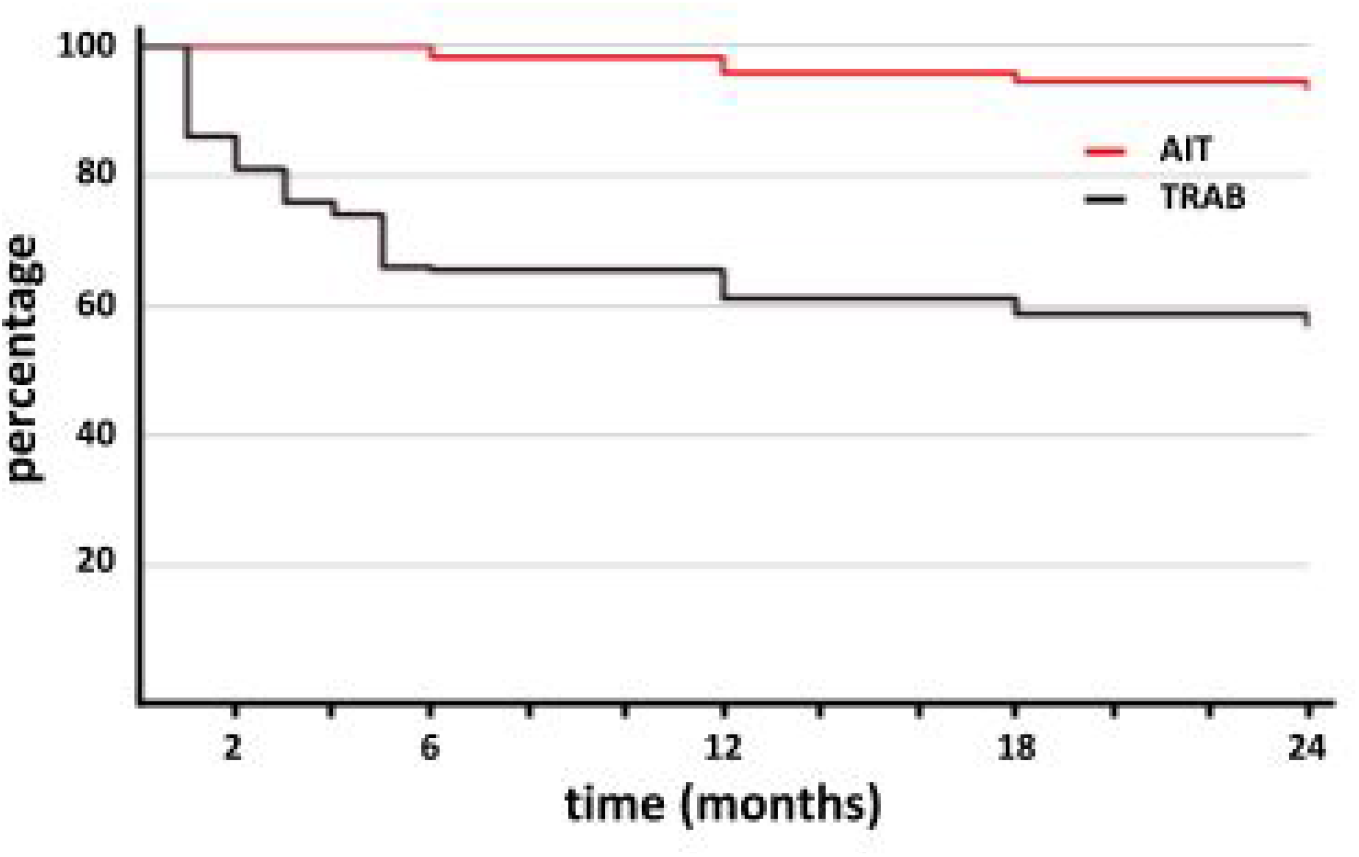
Kaplan-Meier curve indicating how many eyes fulfilled the success criteria (IOP ≤ 21 mm Hg, IOP reduction of at least 20% reduction from baseline, and no secondary surgical interventions including needling in the OR).

## Notes

### Competing Interest Statement

The authors have declared no competing interest.

### Author Declarations

The study protocol was approved by the local ethics committee of the University of Wurzburg, Germany, and performed in accordance with the ethical standards set forth in the 1964 Declaration of Helsinki and the Health Insurance Portability and Accountability Act. Because of its retrospective nature, informed consent was waived.

